# Incidence and associated factors of elderly mortality following hip fracture in Brazil: a systematic review and meta-analysis

**DOI:** 10.1101/2022.02.07.22270577

**Authors:** Viviane Cristina Uliana Peterle, Maria Rita Carvalho Garbi Novaes, Paulo Emiliano Bezerra Junior, Ana Paula Monteiro Gomides Reis, João Carlos Geber Júnior, Amanda Cristina de Souza, Amanda Ribeiro Alves, Natalia Mariana Diogenes Silva de Albuquerque, Júlia Milhomem Mosquéra, Henry Maia Peixoto

## Abstract

**Introduction:** Hip fractures are an important health problem worldwide, and several factors are associated with the mortality. This study aimed to investigate the factors associated with hip fractures in the elderly, based on studies on the population residing in Brazil, and the relationship of fractures with mortality.

**Method:** Prospective and retrospective primary observational studies including hospitalized men and/or women aged 60 or older presenting hip fracture due to bone fragility were selected on the Databases. Independent researchers conducted the study selection process and data extraction. A meta-analysis was performed to determine the hospital mortality rate at 90 days, six months, and one year. The Newcastle-Ottawa scale (NOS) was used to assess the quality of the included studies, and the meta-analysis followed the Preferred Reporting Items for Systematic Reviews and Meta-Analyses (PRISMA).

**Result:** Twenty-five studies totalizing 3,949 patients were included in the systematic review. The population was mainly composed of women (2,680/67.86%). Most patients were in the age group of 70 to 80 years old. Meta-analysis findings: 1) hospital mortality (19 studies, n = 3,175), 10.22% (95% CI 7.27–14.17%; I^2^ 88%); 2) 90-day mortality (3 studies, n = 543), 9.74% (95% CI 3.44–24.62%; I^2^ 90%); 3) six-month mortality (3 studies, n = 205), 24.78% (95% CI 17.07–34.51%; I^2^ 51%); 4) one-year mortality (13 studies, n = 2,790), 21.88% (95% CI 17.5–26.99%; I^2^ 88%). The factors most related to mortality in the studies were: 1) demographic: Older age, male sex; 2) Attributed to clinical conditions: high scoring in preoperative risk scores, comorbidities, neurological/cognitive disorders, functional status; and 3) hospital factors: preoperative period and infections.

**Conclusion:** This review identified variables, including functional status and cognitive changes, related to hip fracture mortality. Knowing these predictors allows for early intervention and planning to adapt health systems to the growing demands of the elderly population.

## Introduction

Hip fractures are an important health problem worldwide and are associated with a decreased quality of life and reduced survival, especially in the elderly population [1]. The risk of short-term mortality is well established, and post-fracture follow-up studies have already shown high mortality within ten years in women and within 20 years in men, indicating the existing relationship between hip fracture and reduced life expectancy [2].

Several studies have investigated the functional results and mortality in elderly patients with hip fractures, identifying outcome predictors and treatment characteristics worldwide [3,4]. However, evidence also associates factors such as a low socio-economic status with a higher risk of death following a hip fracture [5]. In addition to significant differences in age groups and the frequency of fractures, sex and country can also influence mortality [6].

Given the aging of populations worldwide, hip fractures will become a progressively more significant public health burden [7]. Developing countries like Brazil have important and recent demographic changes culminating in an accelerated aging process of their populations [8]. Besides, in the past, acute infectious diseases were the cause of most health problems, while chronic diseases currently significantly impact the population [9].

Official sources of health information confirm this analysis. In the last decade, the number of elderly persons in Brazil increased from 9.31% of the total country population in 2000 to 13.5% in 2018, representing a 45% increase in this age group [10]. In the same period, an increase of 76.9% in hospitalizations due to femur fractures in the Brazilian population over 60 years was observed [11].

A better understanding of the factors associated with hip fractures in the elderly, considering the populations of primary studies on hip fractures carried out in Brazil is required to promote the rational development of intervention strategies, which will may significantly reduce morbimortality morbidity and burdenocio-economic costs. This review aimed to investigate the mortality rate of hip fractures among the elderly in Brazil and its associated factors.

## Materials and methods

This study was registered on the International Prospective Register of Systematic Reviews (Prospero) under number CRD42019129953.

### Eligibility Criteria

Studies with primary source data, classified in their methodology as observational, prospective or retrospective, including case-control and cohort studies, as well as those classified as longitudinal or cross-sectional, which included men and/or women hospitalized for fractured hip (femoral neck, subtrochanteric, intertrochanteric), aged 60 or older, hospitalized due to hip fracture, in study carried out in Brazil.

Studies performed in secondary databases, including article reviews, studies that did not report a relationship with risk factors or that reported in-hospital mortality in patients with acetabular fractures or other femoral fractures not listed above were excluded. In the case of studies with data related to more than one type of fracture, they were eligible only if it was possible to obtain data only for the subgroup of patients with hip fracture. Studies that consider analyzes between independent variables without considering the mortality outcome were also excluded.

Sample size was not part of the exclusion criteria. There was no language or publication date restriction. The minimum age adopted for the elderly population was 60 years, as defined in the Brazilian legislation [12].

The main variables outcomes included were: hospital mortality rates (outcome) and the association of clinical characteristics or other potential risk factors (e.g., age, sex, type of fracture and type of surgery, length of hospital stay, comorbidities, infection, and others). Mortality rates were assessed at 30 days, six months, and one year of follow-up when reported.

### Information sources and search strategy

Relevant studies indexed until March 19, 2019 were searched in the following databases: MEDLINE (via PubMed), Embase, Virtual Health Library (LILACS, IBECS – ES, CUMED – CU, BINACIS – AR, BDENF – Enfermagem, MedCarib, Peru, Lilacs), SciELO, Web of Science, Scopus, Cochrane, and Biblioteca Digital Brasileira de Teses e Dissertações. References from included studies were used as an additional source of data. The search strategy was developed using indexed terms and their synonyms, including terms for “Aged,” “Mortality,” and “Hip Fractures.”

The search strategy was developed using MeSH terms for PubMed, EMTREE terms for Embase, and a combination of keywords, being slightly modified based on each database, as shown in Table 1.

**Table 1.**
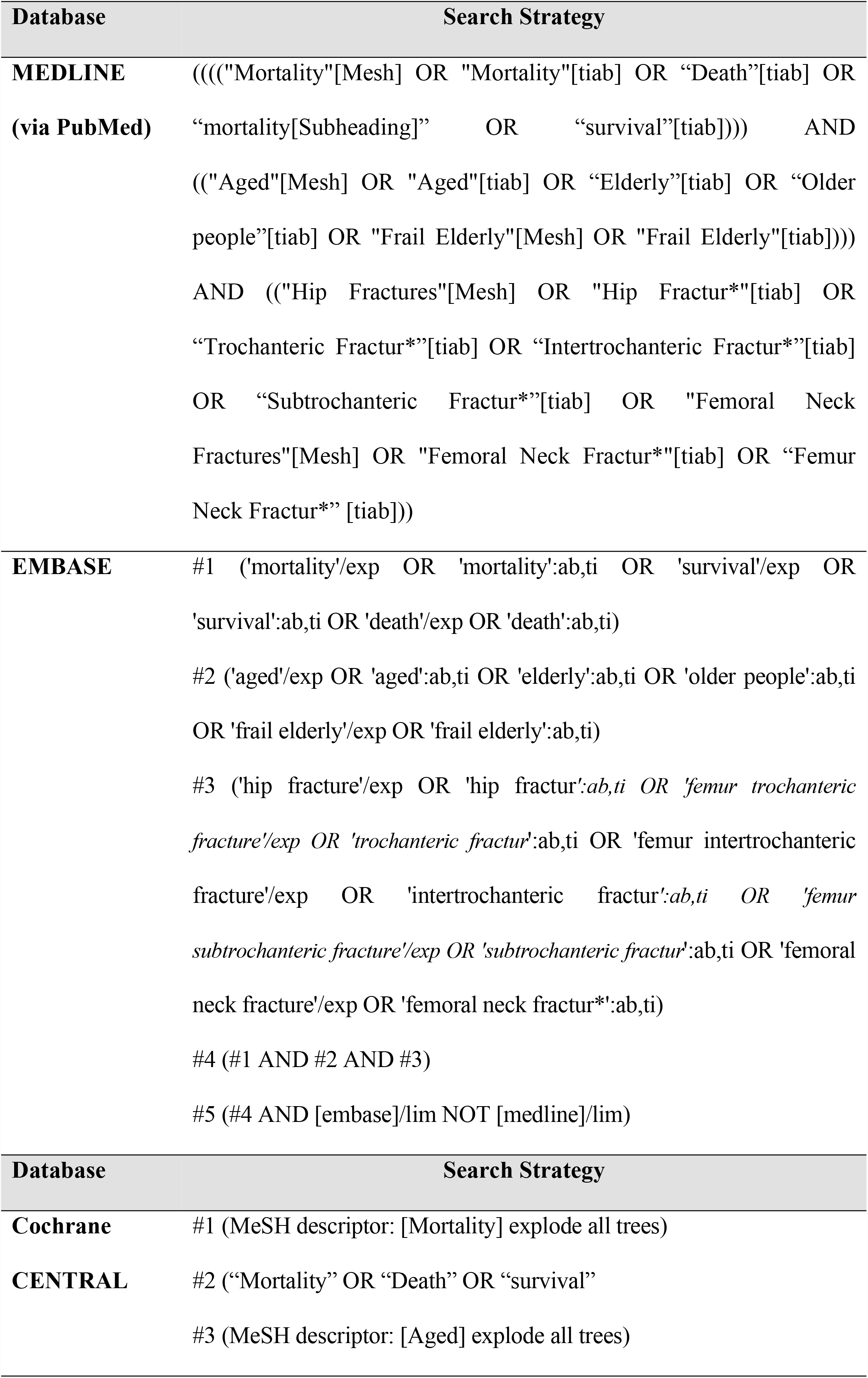
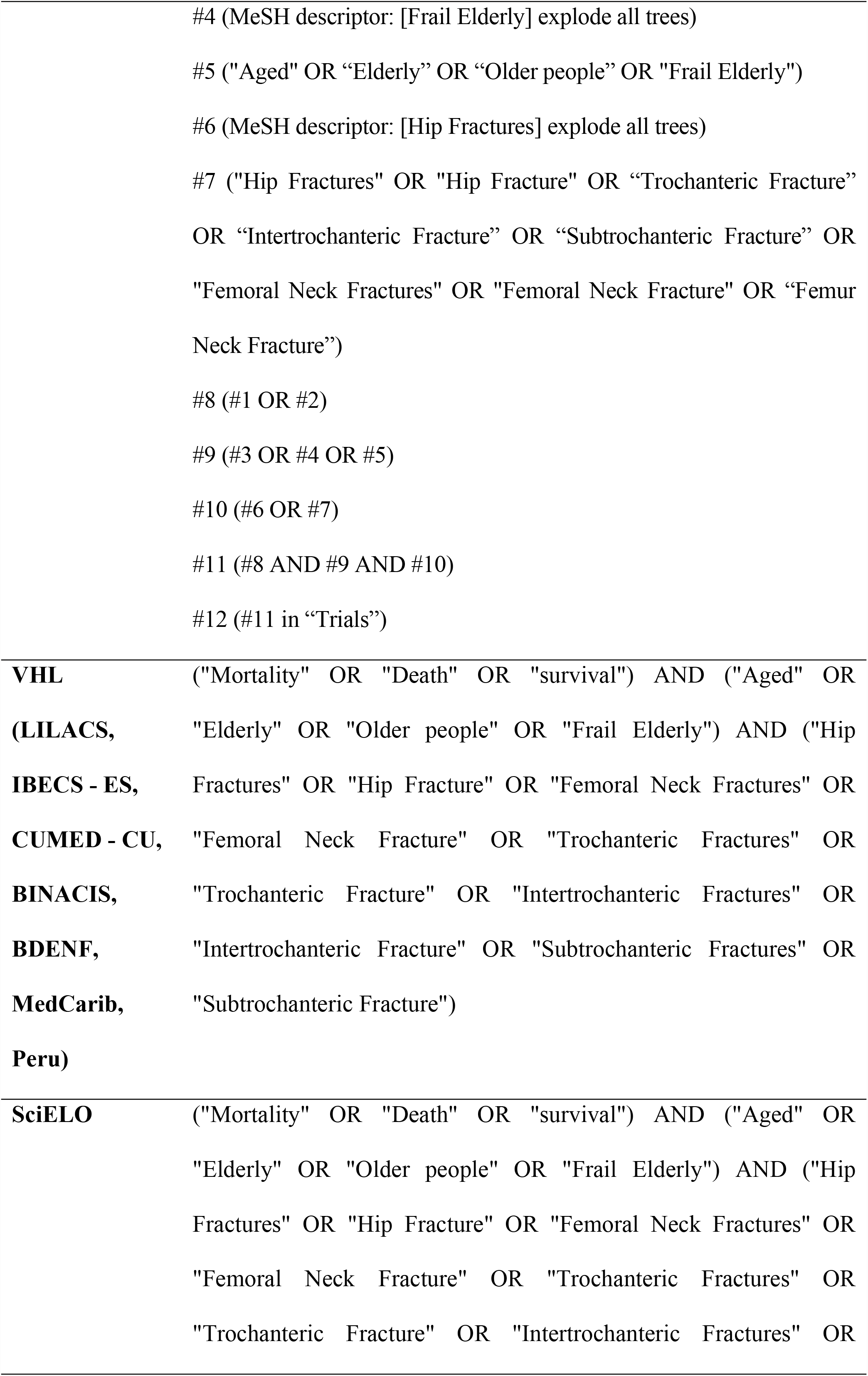
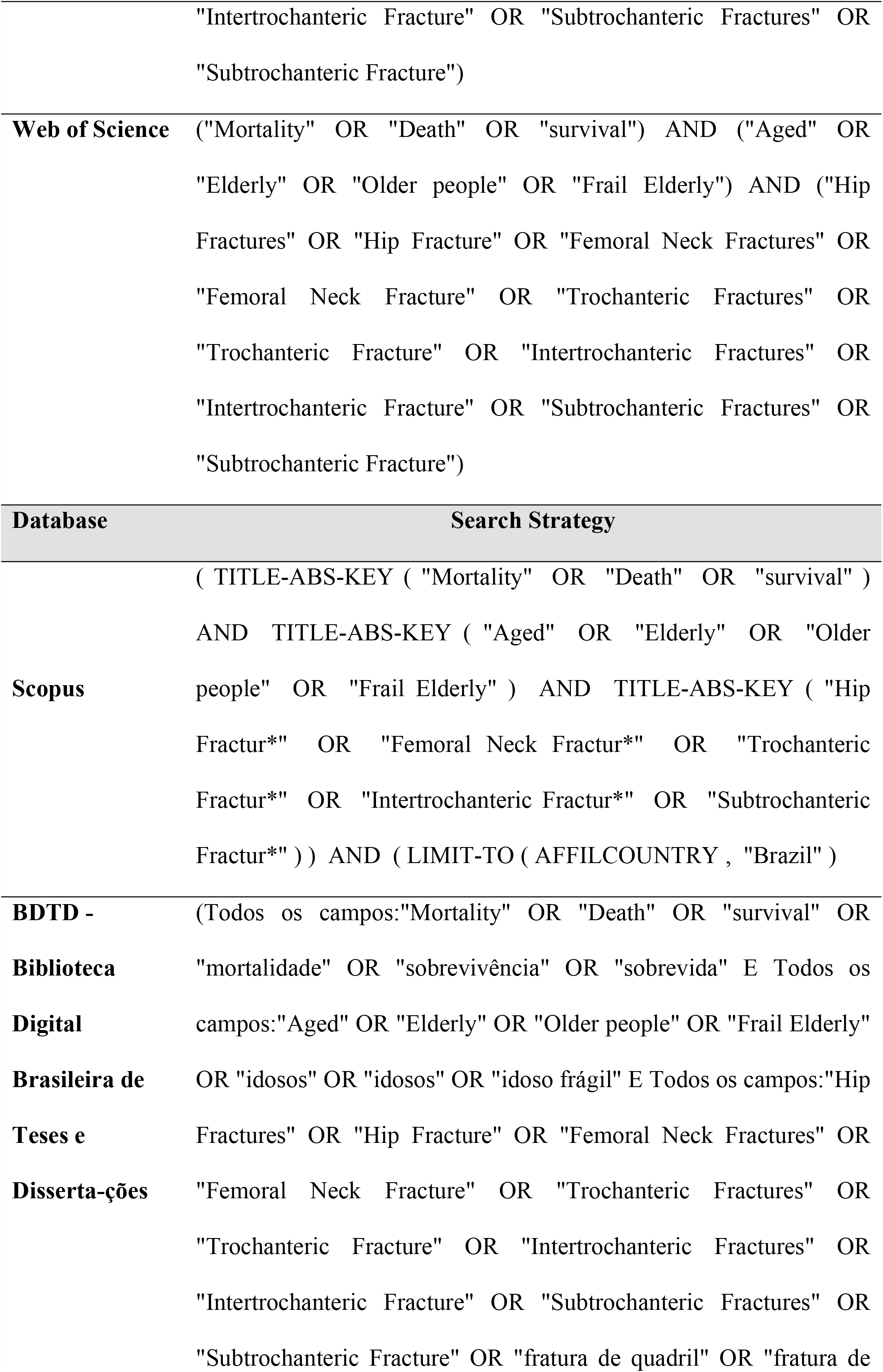
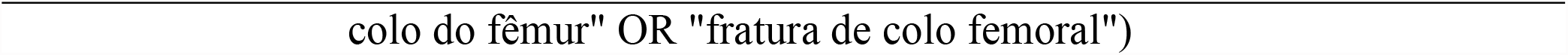
Eletronic search strategies

### Study selection process

The study selection was conducted by four independent researchers (ACS, ARA, JMM, and NMDSA) in two stages. In the first stage, two reviewers independently evaluated each study using a screening checklist based on the abovementioned eligibility criteria. After removing duplicate studies, the pair of reviewers conducted a title and abstract screening, when studies that did not meet the eligibility criteria were excluded.

Potentially and possibly eligible studies were reviewed in the second stage, with the full-text evaluation. If more than one study had the same study population outcomes, the study with the largest sample size or that showed the closest association between risk factors and mortality was selected. Academic conference abstracts were evaluated case-by-case. Disagreements between the pair of researchers on the inclusion of any study were arbitrated by a third researcher (VCUP).

### Data extraction and study quality assessment

The following data were extracted from the selected studies: methodological design, number of patients, characteristics of patients, mean hospital stay, mortality rates, and results of factors associated with mortality as described by the authors of selected studies, which may include association measures such as Odds Ratio (OR), Relative Risk (RR), Hazard Ratio (HR), or *p*-value for association.

Two authors of this study extracted data from the studies separately and independently, with any differences between them being resolved through discussion. If consensus could not be reached, a third author (MRCGN) reviewed the study at issue and arbitrated.

Study quality assessments were performed using the Newcastle-Ottawa Scale (NOS).

### Data analysis

For the quantitative synthesis, the combined mortality rate was estimated through a proportion meta-analysis, employing the inverse of variance method using proportions adjusted by logit transformation. Results were presented as the mortality rate for every 100 patients with the respective 95% confidence interval (CI). Statistical heterogeneity was assessed by the I^2^ inconsistency test, in which values higher than 25% and 50% indicated moderate and high heterogeneity, respectively. Calculations were performed using the random-effects model, which uses the heterogeneity among studies to estimate the CI for the combined measure. All analyses were conducted using the statistical packages meta and metafor in the statistical software R (version 3.6.1) accessed through RStudio software (version 1.2.1335), an integrated development environment for R.

## Results

The literature search identified 9,979 studies. After removing duplicates and evaluating titles, abstracts, and inclusion criteria, 41 of them were submitted to a full-text reading. Of these, 24 studies were included in the systematic review and 19 studies were included in the meta-analysis. Figure 1 details the selection process and the reasons for exclusion.

**Fig 1.**
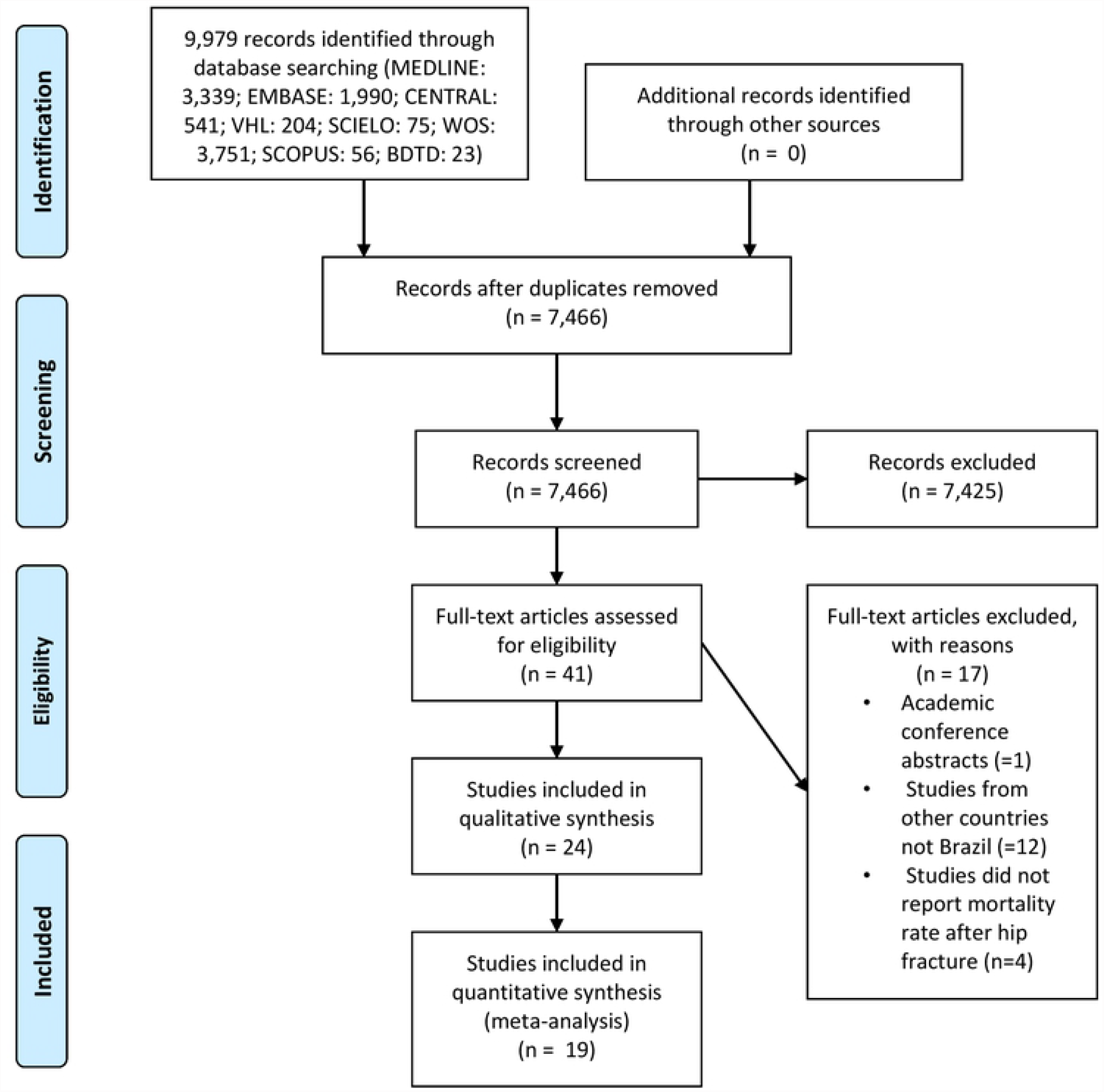
Flow diagram of literature searched and selection criteria

Table 2 shows the characteristics of the studies. A total of 3,949 patients were included in the review. In most articles, the study population consisted predominantly of women, and the proportions of women frequency among them ranged from 84% [13] to 16% [14]. There was a male population predominance in one study, n = 47 (84%) [14].

**Table 2.**
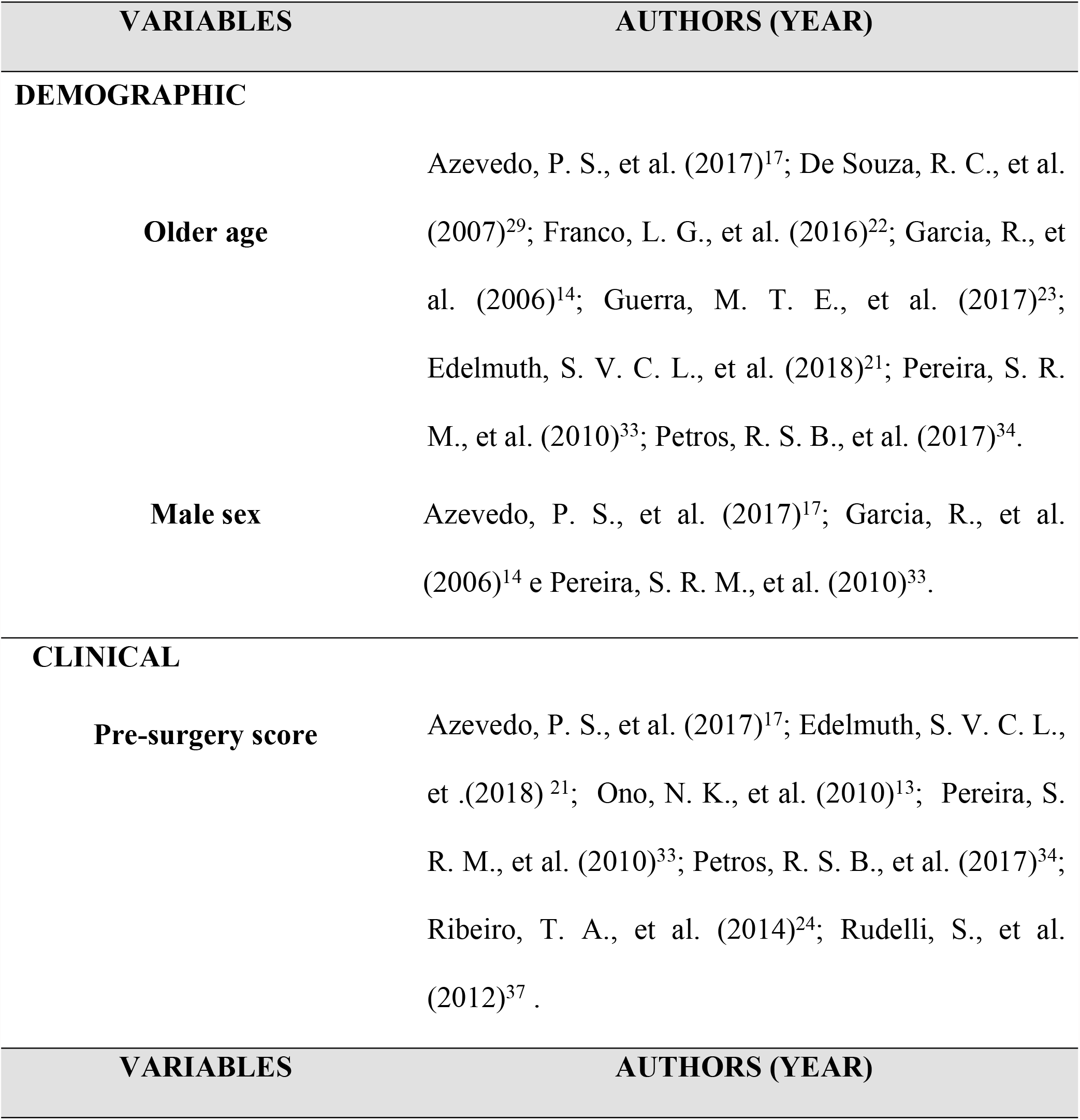
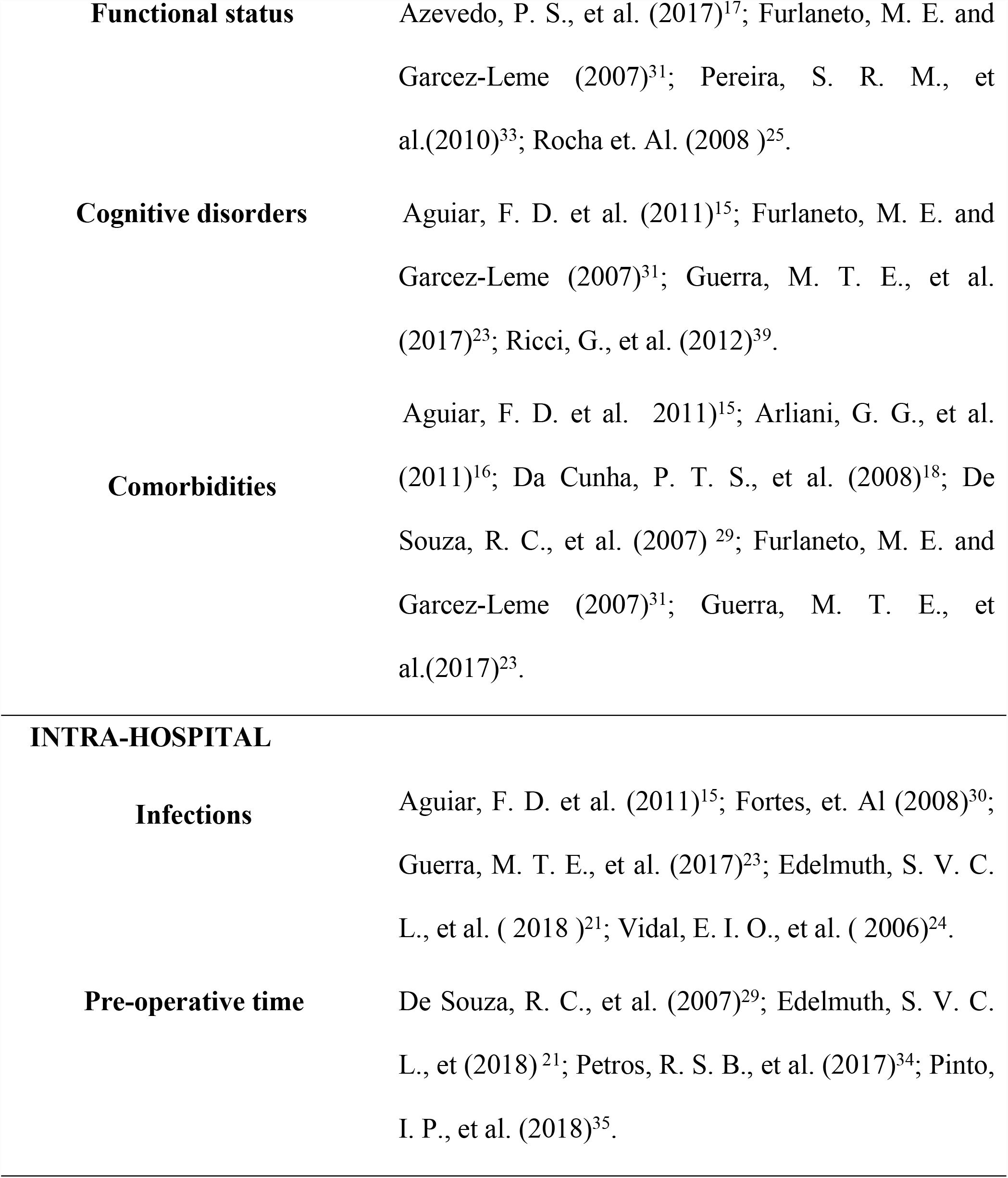
Characteristics of variables analyzed in each study in relation to mortality

| VARIABLES | AUTHORS (YEAR) |
| --- | --- |
| **DEMOGRAPHIC** |  |
| **Older age** | Azevedo, P. S., et al. (2017)<sup>17</sup>; De Souza, R. C., et al. (2007)<sup>29</sup>; Franco, L. G., et al. (2016)<sup>22</sup>; Garcia, R., et al. (2006)<sup>14</sup>; Guerra, M. T. E., et al. (2017)<sup>23</sup>; Edelmuth, S. V. C. L., et al. (2018)<sup>21</sup>; Pereira, S. R. M., et al. (2010)<sup>33</sup>; Petros, R. S. B., et al. (2017)<sup>34</sup>. |
| **Male sex** | Azevedo, P. S., et al. (2017)<sup>17</sup>; Garcia, R., et al. (2006)<sup>14</sup> e Pereira, S. R. M., et al. (2010)<sup>33</sup>. |
| **CLINICAL** |  |
| **Pre-surgery score** | Azevedo, P. S., et al. (2017)<sup>17</sup>; Edelmuth, S. V. C. L., et al. (2018)<sup>21</sup>; Ono, N. K., et al. (2010)<sup>13</sup>; Pereira, S. R. M., et al. (2010)<sup>33</sup>; Petros, R. S. B., et al. (2017)<sup>34</sup>; Ribeiro, T. A., et al. (2014)<sup>24</sup>; Rudelli, S., et al. (2012)<sup>37</sup>. |
| VARIABLES | AUTHORS (YEAR) |
| **Functional status** | Azevedo, P. S., et al. (2017)<sup>17</sup>; Furlaneto, M. E. and Garcez-Leme (2007)<sup>31</sup>; Pereira, S. R. M., et al.(2010)<sup>33</sup>; Rocha et. Al. (2008 )<sup>25</sup>. |
| **Cognitive disorders** | Aguiar, F. D. et al. (2011)<sup>15</sup>; Furlaneto, M. E. and Garcez-Leme (2007)<sup>31</sup>; Guerra, M. T. E., et al. (2017)<sup>23</sup>; Ricci, G., et al. (2012)<sup>39</sup>. |
| **Comorbidities** | Aguiar, F. D. et al. 2011)<sup>15</sup>; Arliani, G. G., et al. (2011)<sup>16</sup>; Da Cunha, P. T. S., et al. (2008)<sup>18</sup>; De Souza, R. C., et al. (2007) <sup>29</sup>; Furlaneto, M. E. and Garcez-Leme (2007)<sup>31</sup>; Guerra, M. T. E., et al.(2017)<sup>23</sup>. |
| INTRA-HOSPITAL |  |
| **Infections** | Aguiar, F. D. et al. (2011)<sup>15</sup>; Fortes, et. Al (2008)<sup>30</sup>; Guerra, M. T. E., et al. (2017)<sup>23</sup>; Edelmuth, S. V. C. L., et al. ( 2018 )<sup>21</sup>; Vidal, E. I. O., et al. ( 2006)<sup>24</sup>. |
| **Pre-operative time** | De Souza, R. C., et al. (2007)<sup>29</sup>; Edelmuth, S. V. C. L., et (2018)<sup>21</sup>; Petros, R. S. B., et al. (2017)<sup>34</sup>; Pinto, I. P., et al. (2018)<sup>35</sup>. |

The predominant age group considering the mean age of patients was 70 to 80 years old [15-27]; one study analyzed nonagenarian patients [28] and one study included the age group from 50 years old on [29].

Regarding geographic regions, 76% of the studies were carried out in the Southeast of Brazil (15-21, 27, 30-36), 12 in São Paulo, six in Rio de Janeiro, and two in Minas Gerais (25, 32). Three studies were carried out in the South of Brazil, and only two with the population from the Northeast region of the country (26, 28, 37). Regarding the time variable, 9 (76%) studies (20-24, 27, 32, 34, 35) considered the mean time between the fracture-related events and death as variables in their analyses. Due to the heterogeneity among studies, the associations were described in Table 2.

Through meta-analysis, mortality rates at 90 days, six months, and one year of hospital stay were estimated in 100 patients. All studies reporting mortality rates for these periods were included in the analysis, except for the study that included only nonagenarians, which was considered too different from the other studies. Only one study reported mortality at 30 days, and the rate was 17.65% [25].

The estimated hospital mortality following hip fracture was 10.22% (95% CI 7.27–14.17%; I^2^ 88%) (Fig 2) considering 19 studies with 3,175 patients analyzed, with high heterogeneity. Only three studies reported mortality at 90 days (n = 543), with an estimated rate of 9.74% (95% CI 3.44–24.62%; I^2^ 90%) (Fig 3), with high heterogeneity. Mortality at six months was also reported only in three studies (n = 205), being estimated at 24.78% (95% CI 17.07–34.51%; I^2^ 51%) (Fig 4). Mortality at one year was reported in 13 studies (n = 2,790) and was estimated at 21.88% (95% CI 17.5– 26.99%; I^2^ 88%) (Fig 5). Additionally, one study had a longer follow-up period, with mortality at 24 and 48 months of 26% and 40% in patients without delirium and of 36% and 60% in patients with delirium during hospitalization after a fracture [31].

**Fig 2.**
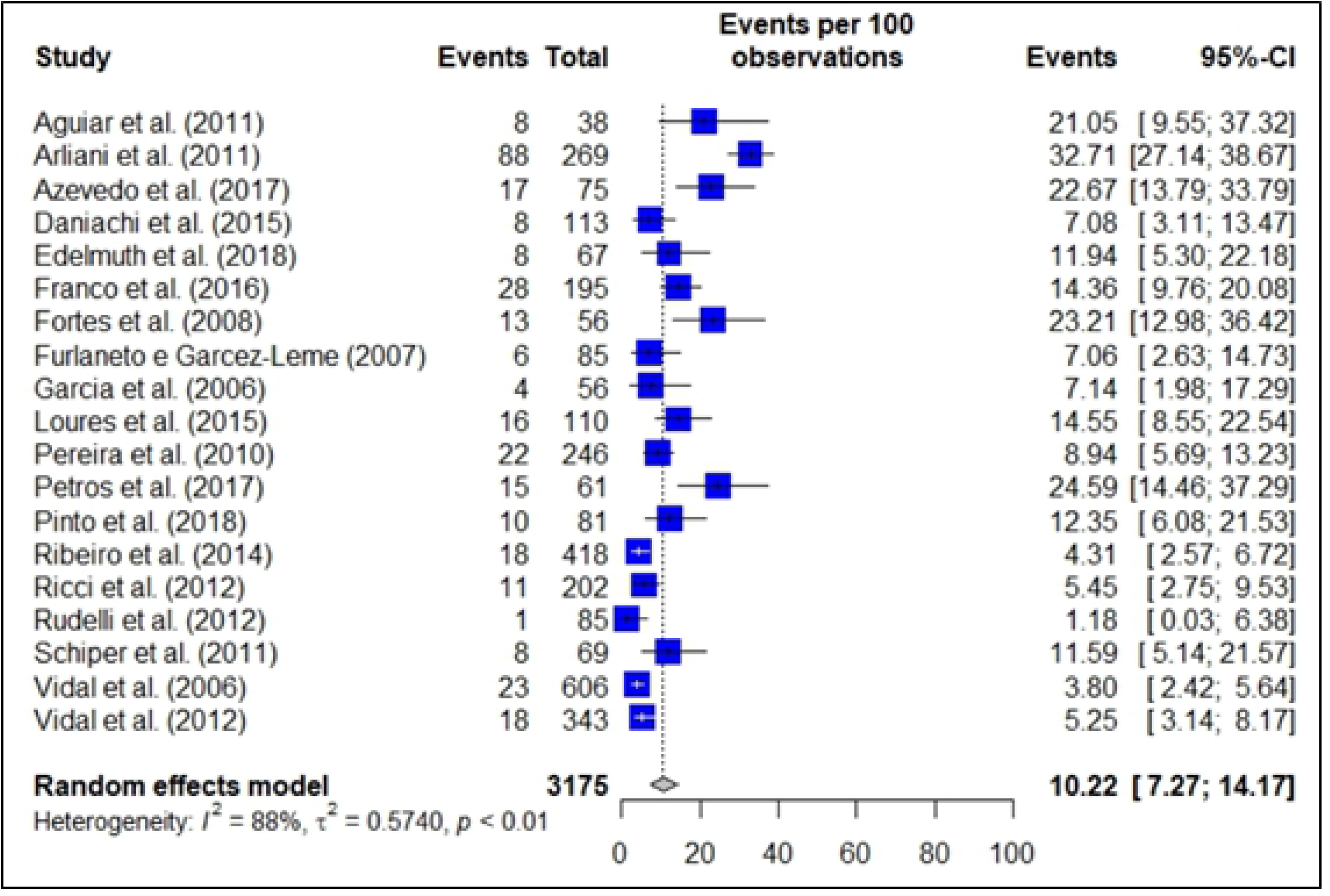
Forest-plot for hospital mortality following hip fracture (n= 3,175)

**Fig 3.**
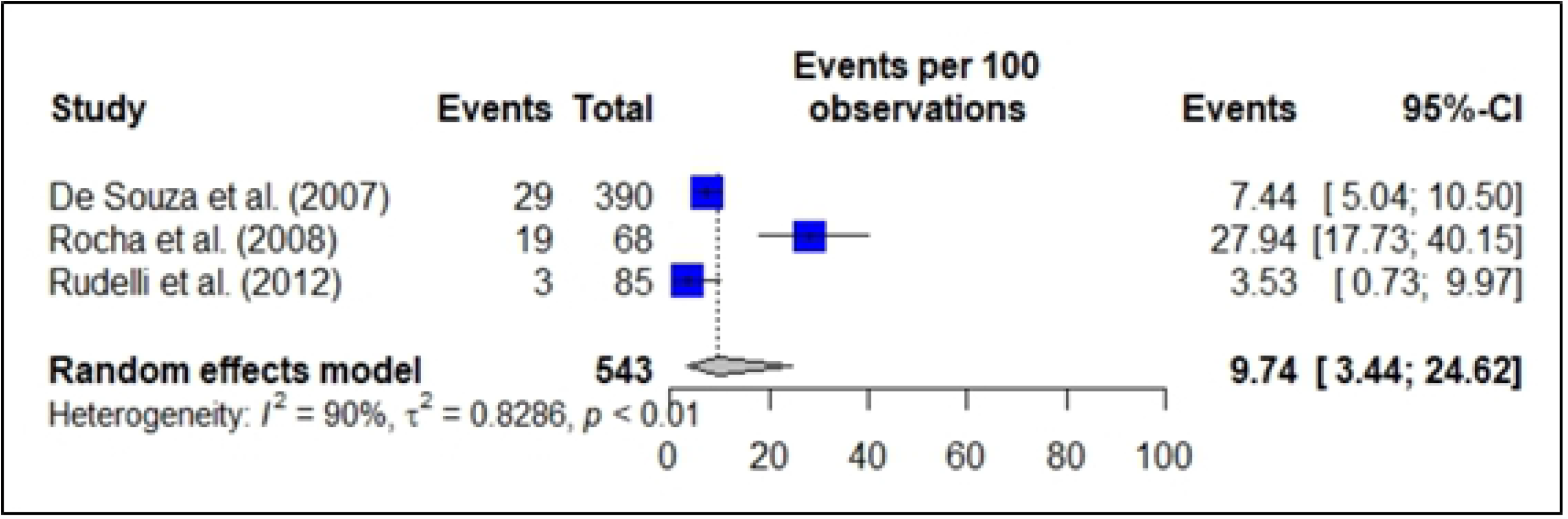
Forest-plot for hospital mortality following hip fracture at 90 days (n = 543)

**Fig 4.**
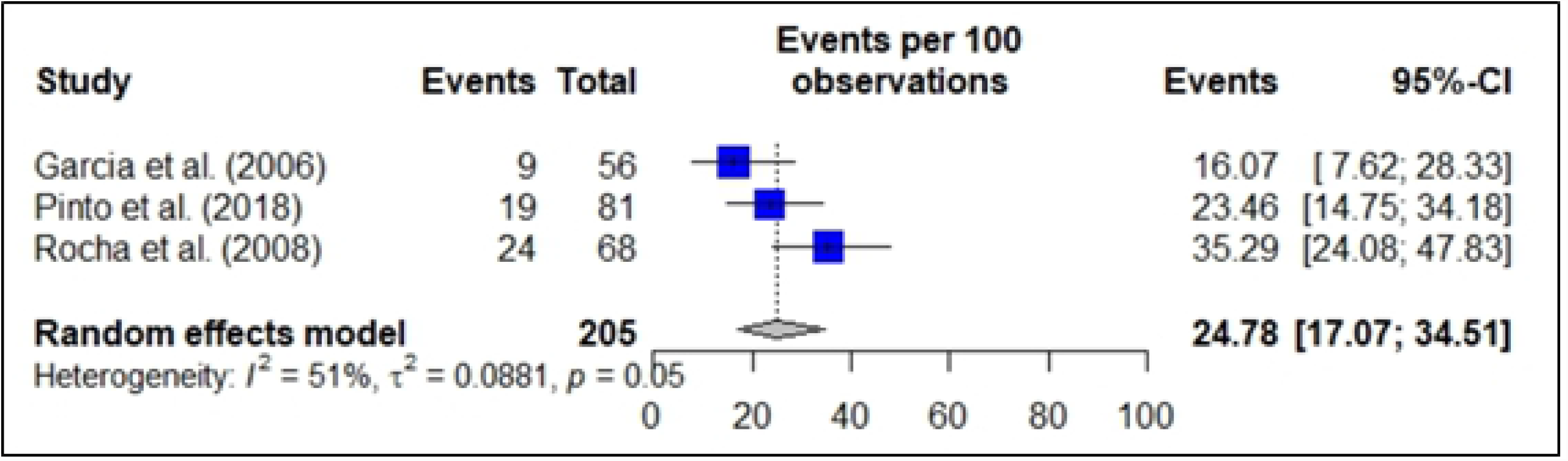
Forest-plot for hospital mortality following hip fracture at 6 months (n = 205)

**Fig 5.**
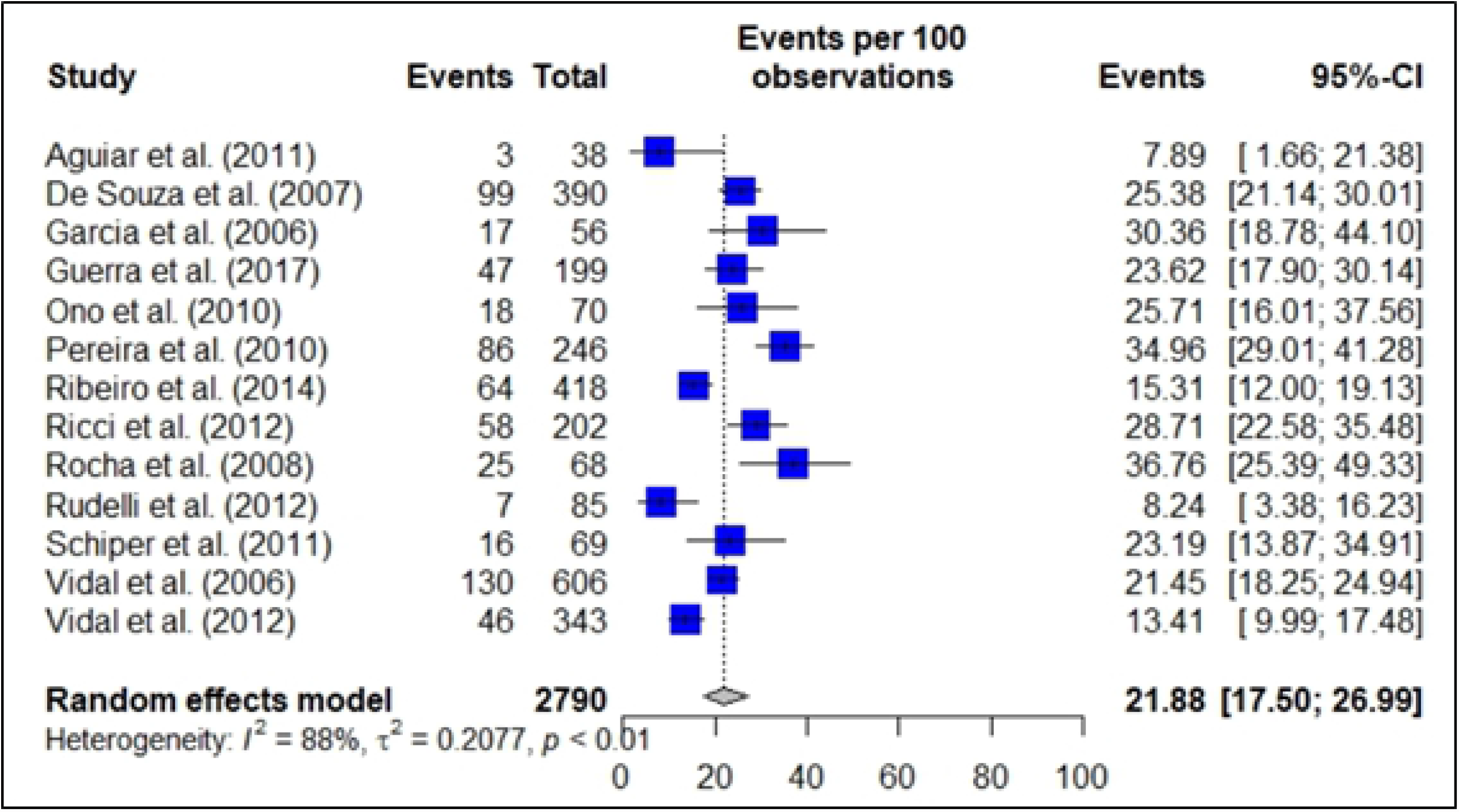
Forest-plot for hospital mortality following hip fracture at one year (n= 2,790)

Due to the heterogeneity among the variables analyzed in each study regarding the factors associated with mortality following femur fracture in the elderly in Brazil, the predictors identified in this systematic review were grouped into 1) demographic factors: older age [14,17,20-23,33,34] and male sex [14,17,33]; 2) clinical factors: comorbidities [15,16,18,20,23,31], high scoring in surgical risk assessment scores [13,17,21,24,33,34,38], functional status before fracture [17,31,33,36], and cognitive/neurological disorders [15,23,31,39]; and 3) intra-hospital factors: infections [15,21,23,26,30] and the time between hospitalization and surgery [20,21,34,35]. Only one study addressed prolonged hospitalization as a factor associated with mortality [16] (S1Table).

## Discussion

Despite the limitation in the numerical data extraction due to the great variability of studies and to the number of variables analyzed in each study, the review aimed to synthesize the information generated by traumatology and orthopedics services in Brazil in the management of hip fractures over the years and to compare it with the international literature considering the factors associated with the elderly mortality.

The Brazilian National Public Health System has already considered the hospitalization rate for femur fracture in Brazil as an indirect marker of the quality of care for the elderly population [40]. It was due to the correlation between fractures and the high mortality [1], representing the result of a series of health processes, from primary fall prevention in primary healthcare to hospital and post-discharge rehabilitation strategies [26].

The relationship between aging and comorbidities [41] and comorbidities associated with high mortality following hip fractures are the object of several studies [42,43]. The predominance of long-lived women in the hospitalization profile found in the synthesis of fracture care services in Brazil reinforces the osteoporosis association as a baseline disease. Worldwide, hip fractures can be considered the main complication of mortality and economic burden secondary to osteoporotic disorder [44].

Several studies have reported the postoperative mortality rate after surgical treatment for hip fracture [45]. The variation in mortality rates among the services analyzed also reflects the variation in mortality among the world population [46]. In Brazil, there are scarce accurate data available on the prevalence of osteoporosis, the incidence of falls and fractures, and the costs related to these events [33]. This can be seen in the wide variation in methodologies and variables among the included studies.

The common evidence among Brazilian studies regarding the variables male sex, aging, and comorbidity agrees with the world literature, and all of them are related to increased mortality [47].

The concern of Brazilian studies in evaluating other variables involved and associated with mortality, such as cognitive disorders and functional deficits, is noteworthy. Several studies have investigated the relationship between these variables and mortality in elderly patients following hip fractures, identifying the predictor factors of the patient outcome and the treatment characteristics [48,49].

A better understanding of the factors involved in fracture consolidation is necessary to promote the rational development of preventive therapies, which will significantly reduce morbidity and socio-economic costs [50].

Recommendations already evaluated, such as nutritional intake optimization, rehabilitation programs (at home), and the possibility of having psychological counseling in patients with difficulties in psychosocial dimensions, also apply after hip fracture surgery [51].

An important limitation found in the studies was the time factor. Despite many studies have included this relationship as an analysis variable, related biases, such as public versus private service, the time between fracture and access to traumatology and orthopedics services, conditions to timely provide adequate surgical treatment, clinical conditions of patients, risk prediction for decision making, multi-professional support regarding nutritional, respiratory, and rehabilitation factors, and support for caregivers, sometimes cannot be included in the analyses, but are present as non-measurable associated factors affecting mortality rates over time.

## Conclusion

Hip fractures are associated with a high morbimortality burden. Studies on the variables associated with hip fractures in Brazil reflect the country’s growing population aging, permeated by multiple comorbidities, with osteoporosis as the baseline disease. The heterogeneity among short-term mortality rates reflects the non-measurable factors related to the management of fractures, while the high incidence of mortality in the long-term post-fracture period shows the impact of the event as a milestone in the life history of patients. Therefore, the studies included in this research throw some light on the quality of health services, as they establish, among others, time and outcome indicators that require improvement efforts. Initiatives to increase quality in managing patients with hip fracture are important due to the high burden of the population morbimortality. Differences among studies regarding the pre- and postoperative time also suggest a subjectivity in managing fractures, such as in the access to health services, ability to timely perform the required procedure, and hospital support in the pathology management.

## Data Availability

All relevant data are within the manuscript and its Supporting Information files.

## SUPPORTING INFORMATION TITLES

**S1 Table**. Characteristics of included studies and mortality following hip fracture.

